# Cerebrospinal fluid D-2-hydroxyglutarate for IDH-mutant glioma: utility for detection versus monitoring

**DOI:** 10.1101/2025.04.08.25325500

**Authors:** Cecile Riviere-Cazaux, Yuta Suzuki, Zain Kizilbash, William J. Laxen, Jean M. Lacey, Tamara M. Wipplinger, Arthur E. Warrington, Michael B. Keough, Lionel Fotso Kamga, Katherine M. Andersen, Nicholas Canaday, Matthew L. Kosel, Silvia Tortorelli, Ugur Sener, Michael W. Ruff, Paul A. Decker, Jeanette E. Eckel-Passow, Sani H. Kizilbash, Timothy J. Kaufmann, Terry C. Burns

## Abstract

**BACKGROUND:** Imaging-based monitoring of gliomas is limited by treatment-related changes. D-2-hydroxyglutarate (D-2-HG), produced by the isocitrate dehydrogenase (IDH) mutation, is detectable in cerebrospinal fluid (CSF) that can be accessed from various anatomic compartments. We evaluated CSF D-2-HG as a serially accessible biomarker for IDH-mutant gliomas.

**METHODS:** A CLIA-approved gas chromatography mass spectrometry assay was developed for CSF D– and L-2-HG. Lumbar and cranial CSF samples were collected from patients with IDH-mutant gliomas or IDH-wild-type brain tumors and non-tumor pathologies via surgical field collection, lumbar punctures, Ommaya reservoirs, and ventriculoperitoneal shunts.

**RESULTS:** CSF D-2-HG was significantly higher in cranial than lumbar samples from IDH-mutant glioma patients (median lumbar=0.20 μM, cranial = 1.72 μM; p<0.0001). Cranial, but not lumbar, CSF D-2-HG distinguished primary IDH-mutant gliomas from IDH-wild type lesions (cranial AUC= 0.89, 95% confidence interval (CI)= 0.80-0.97); lumbar AUC= 0.52, 95% CI=0.28-0.76). When evaluated in recurrent lesions as a separate validation cohort, this finding was also reproduced in this group (cranial AUC=0.97, 95% CI= 0.94-1.00; lumbar AUC=0.60, 95% CI=0.38-0.83). Cranial CSF D-2-HG levels decreased to 0.54x of baseline with resection in seventeen patients (p=0.0129) but did not decrease significantly with chemoradiation in five patients (p=0.6250). Longitudinal anatomical changes, such as cavity collapse, influenced serial sample interpretation. In grade 4 IDH-mutant astrocytomas, serial cranial CSF D-2-HG increased with disease progression and differentiated stability from pseudoprogression when tumor-CSF contact was sufficient.

**CONCLUSIONS:** Serial cranial CSF D-2-HG shows promise as a monitoring biomarker in patients with IDH-mutant gliomas when anatomic variables remain constant.

**KEY POINTS:**

1. Cranial CSF D-2-HG levels exceed that of lumbar CSF in patients with IDH-mutant gliomas.
2. Cranial CSF D-2-HG may discriminate disease stability vs. treatment effects, although post-resection anatomical changes can impact monitoring.

**IMPORTANCE OF THE STUDY:** Improved glioma monitoring is needed due to challenges distinguishing disease progression from treatment-related changes on imaging. Toward this goal, we evaluated CSF D-2-HG as a biomarker of IDH-mutant gliomas using a CLIA-approved assay. This study answers whether D-2-HG can identify IDH-mutant gliomas via either cranial or lumbar CSF. Importantly, in seventeen patients, we demonstrate that CSF D-2-HG is responsive to cytoreduction via resection, but not chemoradiation in five patients. This is also the first study to demonstrate that longitudinal anatomical changes can impact evaluation of CSF D-2-HG as a monitoring biomarker. Finally, the study demonstrates that serial CSF D-2-HG can increase with disease progression, but not pseudoprogression or stable disease, in five patients with grade 4 IDH-mutant astrocytomas. These findings support the potential of CSF D-2-HG as a monitoring biomarker in patients with IDH-mutant gliomas, particularly when there are minimal changes to the anatomy of the resection cavity.

## INTRODUCTION

Isocitrate dehydrogenase (IDH)-mutant astrocytomas and oligodendrogliomas are the most common gliomas in young adults^1^. Although these tumors have a better prognosis compared to their IDH-wild type counterparts, they inevitably progress to higher-grade, fatal lesions. While vorasidenib, an IDH inhibitor, was recently approved for grade 2 IDH-mutant gliomas^2^, treatment options for high-grade IDH-mutant gliomas (HGGs) remain limited. Glioma monitoring using two-dimensional cross measurements via Response Assessment in Neuro-Oncology (RANO 2.0) criteria is hindered by the insidious progression of low-grade gliomas and post-radiation increases in contrast-enhancement that mimic disease progression^3^, underscoring the need for improved response assessment tools.

IDH mutations result in increased production of D-2-hydroxyglutarate (D-2-HG)^4,5^, a stable and quantifiable oncometabolite that serves as a candidate biomarker. While magnetic resonance spectroscopy has been available for over a decade to measure total 2-HG^5–7^, technical challenges have limited widespread adoption. Evaluation of D-2-HG in liquid biopsies, particularly cerebrospinal fluid (CSF), may offer a more practical alternative in clinical settings. Unlike plasma, where sensitivity is reduced due to a low signal-to-noise ratio^8^, CSF is proximal to the tumor and has been reported to contain elevated D-2-HG concentrations in IDH-mutant gliomas compared to IDH-wild type controls when analyzed via mass spectrometry^9–11^.

Despite this potential, few studies have rigorously evaluated the diagnostic performance of CSF D-2-HG^10,11^ or its responsiveness to changes in disease burden. Our prior work included initial findings from six patients with CSF D-2-HG measured before and after resection using CSF access devices^9^. Using our Clinical Laboratory Improvement Amendments (CLIA)-certified assay for CSF D-and-L-2-HG, we assess the diagnostic utility of CSF D-2-HG in lumbar and intracranial CSF, as well as its variability across CSF compartments within patients. Additionally, we evaluate the effects of resection and chemoradiation on CSF D-2-HG levels in IDH-mutant gliomas. Finally, we present longitudinal CSF data from patients with grade 4 IDH-mutant astrocytomas to understand its potential for assessing therapeutic response and disease trajectory.

## MATERIALS AND METHODS

### Patient recruitment to research studies

Patients were recruited and informed consent obtained for one or more Institutional Review Board-approved protocols. The Mayo Clinic Neuro-oncology biorepository allows CSF collection from the surgical field and provides access to CSF collected for clinically indicated reasons, or under other protocols. A subset of patients underwent Ommaya reservoir placement at the time of resection (NCT04692337) or biopsy (NCT06322602) to enable longitudinal CSF access for biomarker discovery. A separate liquid biopsy protocol (NCT04692324) was used to bank CSF samples acquired via lumbar puncture and/or from sampling of existing clinically indicated CSF access devices, such as ventriculoperitoneal shunts or external ventricular drains. The Mayo Clinic neuro-oncology biorepository approved data use across studies under IRB 17-004675.

### CSF collection and Ommaya reservoir placement

Most intracranial CSF samples were obtained intra-operatively at the time of resection. CSF was sampled from the subarachnoid space (most often an exposed sulcus or cistern after dural opening), a prior resection cavity, a tumor cyst and/or the lateral ventricle if surgically accessed. In patients electing to undergo Ommaya reservoir placement at resection, catheters were placed in the resection cavity after tumor resection. Some, but not all, cavities were continuous with the ventricular system. A separate incision was used for reservoirs placed at biopsy. Our study protocols were previously reported and are available online^12^. The most frequently utilized Ommaya reservoir had a 1.5 cm side with a flat bottom and side inlet (Natus ® NT8501214), which was connected to an antibiotic-coated ventricular catheter (Codman ® Hakim Bactiseal Catheter, NS5048). CSF samples were acquired at the time of magnetic resonance imaging. CSF was centrifuged at 400xG for 10 minutes at 4°C, aliquoted, and stored at –80 ° C. De-identified patient-level clinical and technical metadata are provided in **Supplementary Data**.

### D-and-L-2-HG quantification and CLIA validation

D-and-L-hydroxyglutarate were quantified in CSF using a CLIA-level gas chromatography-mass spectrometry (GCMS)-based assay in the Mayo Clinic Biochemical Genetics Laboratory. To meet CLIA guidelines for clinical deployment of the assay, intra-and-inter-assay precision, accuracy (recovery), reportable range and linearity, and analytical specificity (or assay interference) were measured. Please see *Supplemental materials* for detailed methods, as well as results of the impact of different temperatures or exposure to whole blood or plasma on CSF D-and-L-2-HG abundance, and results confirming absence of assay interference by agents that may be used within the surgical field, including antibiotics (**Supplementary Figure 1**). Beginning in 2023, for increased consistency across batches, three stock CSF samples were run in each batch (0.5, 5, and 50 μM) to generate calibration curves, the slopes of which were used to adjust CSF D-or-L-2-HG values for that run based on expected versus measured values.

### Statistical analyses

For temperature and blood experiments in Supplemental Figure 1, one-way ANOVA with Tukey’s multiple comparison test were performed; an unpaired two-tailed t-test was performed for the antibiotic experiment. For each analysis in Figure 1 onward, non-normal distribution of D-2-HG was identified by D’Agostino-Pearson test. As such, unpaired analyses were performed by Mann Whitney U-tests and paired tests by Wilcoxon-signed rank tests. Receiver operating characteristic (ROC) curves were generated, and area under the curve (AUC) analyses were performed, with a 95% confidence intervals obtained using the Wilson/Brown method. Optimal sensitivity and specificity for the cut-off were identified by calculating the Youden index as (sensitivity + specificity)-1 at each unique value in the dataset. Graphs and statistical analyses were generated using GraphPad PRISM 10.3.0.

**Figure 1.**
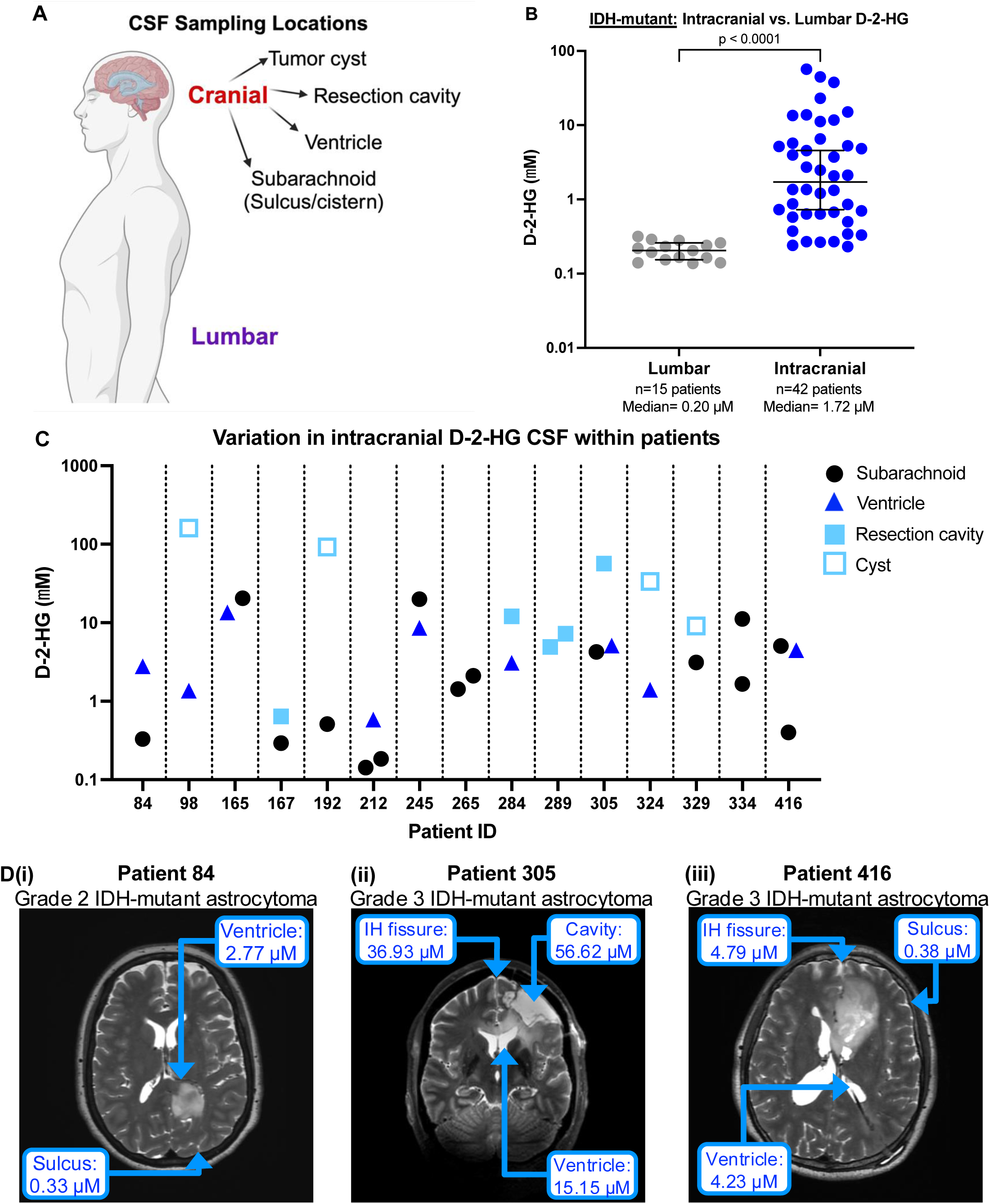
D-2-hydroxyglutarate concentrations vary across cerebrospinal fluid compartments. **(A)** CSF can be sampled from lumbar or cranial compartments, with cranial fluid including tumor cystic fluid, resection cavity fluid, and ventricular or subarachnoid CSF. **(B)** CSF D-2-HG concentrations in cranial (n=42 patients) or lumbar (n=15 patients) were compared via Mann-Whitney U-test. The line is at the median, with a 95% confidence interval. **(C)** CSF D-2-HG concentrations in the 15 patients wherein at least 2 cranial CSF samples were obtained from different locations during surgery. **(D)** Three example patients are shown from (C), demonstrating CSF D-2-HG concentrations in at least two compartments from (i) Patient 84, (ii) Patient 305, and (iii) Patient 416.

## RESULTS

### Anatomical variation in CSF D-2-HG abundance

In total, 417 CSF samples to date have been quantified for CSF D-or-L-2-HG from 194 unique patients, 77 of whom had an IDH-mutant lesion (17 grade 2, 21 grade 3, 1 grade 2 to 3, and 16 grade 4 IDH-mutant astrocytomas; 9 grade 2 and 13 grade 3 oligodendrogliomas). 107 patients had known IDH-wild type pathologies, including glioblastoma (n=54 patients), metastatic brain tumors (n=20 patients), and a variety of other pathologies including but not limited to acoustic neuromas, other diffuse gliomas (not otherwise specified), ependymomas, and gangliogliomas. The remaining ten patients had an unknown IDH status (n=9 patients) or a variant of unknown significance (n=1 patient). 169 CSF samples originated from the surgical field, 116 from lumbar punctures, 120 from CSF access devices, and 12 from EVDs or other locations (e.g., drained pseudomeningoceles).

CSF can be sampled from the lumbar cistern or from various cranial locations (**Figure 1A**). We previously reported a significant difference in D-2-HG from paired intracranial-versus-lumbar CSF samples^12^. To determine whether this difference held in a larger cohort of unpaired samples, we compared CSF D-2-HG from cranial or lumbar CSF samples in primary tumors or controls. As with paired samples, CSF D-2-HG was significantly higher in cranial (n=42) than lumbar (n=15) IDH-mutant glioma CSF samples (median lumbar=0.20 μM, cranial=1.72 uM; p<0.0001) (**Figure 1B**). This result held even when evaluated in a separate patient cohort where samples were obtained after prior resection during latent or recurrent disease (median lumbar=0.19 μM, cranial=0.71 μM; n=13 versus 22 patients, respectively; p<0.0001) (**Supplementary Figure 2A**). Interestingly, intracranial CSF D-2-HG was also significantly higher than lumbar D-2-HG in IDH-wild type tumors or non-tumor pathologies, although the magnitude of the cranial-versus-lumbar fold-change was much lower than in IDH-mutant gliomas (median lumbar=0.17 μM, cranial=0.24 μM; p<0.0001) (**Supplementary Figure 2B**).

In addition to variation between cranial versus lumbar compartments, cranial CSF 2-HG concentrations can vary markedly within the cranial compartment^12^. As such, we evaluated D-2-HG concentrations in CSF sampled from at least two different intracranial locations per patient, when able. In fifteen IDH-mutant glioma resections, the median CSF D-2-HG fold-change within a patient was 4.07x (range: 1.47-180.89x) when evaluating the lowest versus highest D-2-HG concentration (**Figure 1C**). Three examples of patients are shown in Figure 1D, demonstrating low CSF D-2-HG concentrations in sulcal CSF not in direct contact with the tumor (**Figure 1Di, iii**) and elevated CSF D-2-HG in resection cavity fluid (**Figure 1Dii**). Indeed, for patients with multiple samples including a resection cavity (n=4), the resection cavity sample had the highest CSF D-2-HG level in all 4 cases (**Figure 1C**). Such findings in resection cavity fluid could contribute to the trend toward higher levels of CSF D-2-HG in recurrent IDH-mutant gliomas, wherein eight of the nineteen recurrent CSF samples were obtained from a resection cavity (median primary=1.325 μM, recurrent=4.630 μM; p=0.2034) (**Supplementary Figure 3**). Finally, CSF D-2-HG concentrations varied across patients and had a low R^2^ of 0.3009 when correlated with tumor volume (**Supplementary Figure 4A)**, consistent with previously reported variations in D-2-HG production per cell across patients^13^, as well as individualized anatomic variables. In summary, CSF D-2-HG is higher in cranial than lumbar CSF, but D-2-HG concentrations also vary across cranial compartments, even within a patient.

### CSF D-2-HG for discriminating IDH-mutant versus wild-type glioma

We then asked whether CSF D-2-HG could discriminate an IDH-mutant glioma from an IDH-wild type lesion. Given differences in lumbar versus cranial CSF D-2-HG concentrations, comparisons between IDH-mutant gliomas and IDH-wild type samples were performed separately for each location. To test these findings in two separate cohorts, the cohort was divided into primary versus recurrent lesions and analyses performed separately in each group. We first evaluated banked cranial CSF from patients with primary lesions with no prior treatment. Seventy-three samples were evaluated, of whom 35 patients were determined to have IDH-mutant gliomas, and 38 patients had IDH-wild type pathologies (n=27 gliomas, n=11 other brain tumors or non-tumor pathologies). Receiver operating characteristic (ROC) curves demonstrated that cranial CSF D-2-HG concentration can discriminate IDH-mutant gliomas from IDH-wild type samples with an area under the curve of 0.89 (95% confidence interval: 0.80-0.97) (**Figure 2Ai**). Using the Youden index, 0.33 μM was identified as the CSF D-2-HG cut-off with the optimal balance between sensitivity and specificity (80% and 89.47%, respectively). A Mann Whitney U-test confirmed significantly higher intracranial CSF D-2-HG concentrations in primary IDH-mutant gliomas than IDH-wild type samples (median IDH-mutant=1.33 μM, IDH-wild type=0.23 μM; p<0.0001) (**Figure 2Aii**). By comparison, lumbar CSF D-2-HG poorly distinguished primary IDH-mutant gliomas from IDH-wild type samples at an AUC of 0.52 (n=13 patients each; 95% CI: 0.28 to 0.76) (**Figure 2Bi**). Consistent with this finding, lumbar CSF D-2-HG was not significantly different between IDH-mutant gliomas versus IDH-wild type samples (median IDH-mutant=0.14 μM, IDH-wild type=0.17 μM; p=0.8695) (**Figure 2Bii**).

**Figure 2.**
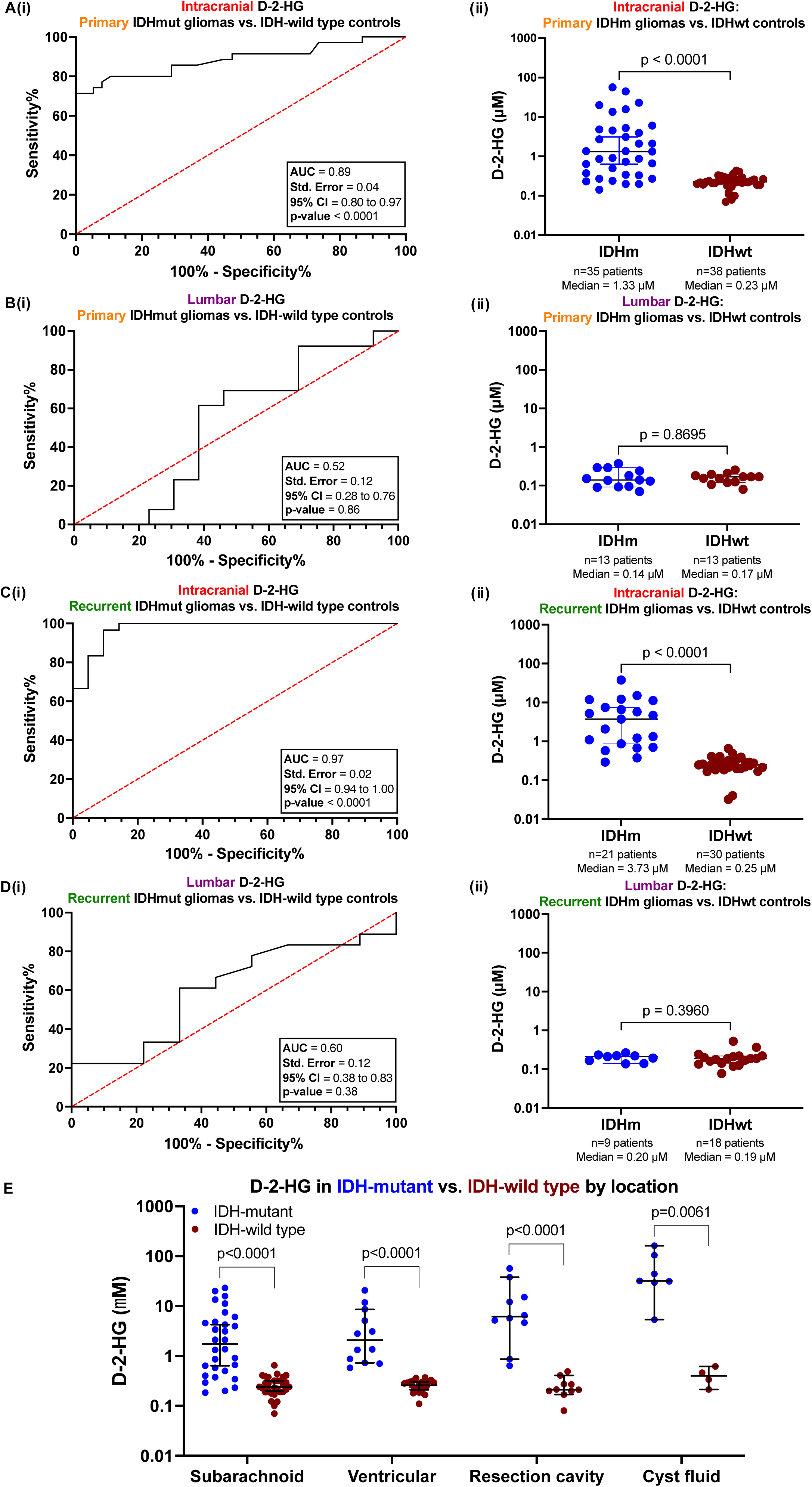
CSF D-2-HG can distinguish cranial, but not lumbar, IDH-mutant gliomas from IDH-wild type samples. (A) **(i)** Receiver operating characteristic (ROC) analyses were performed to evaluate the sensitivity versus specificity of different cranial CSF D-2-HG for distinguishing primary IDH-mutant gliomas from IDH-wild type samples (n=35 versus 38 patients, respectively). **(ii)** A Mann-Whitney U test was also performed on these patients’ samples. The line is at the median, with bars indicating the 95% confidence interval. Similar analyses were performed in **(B)** lumbar CSF from primary IDH-mutant gliomas versus wild type samples (n=13 patients per group), **(C)** cranial CSF from recurrent IDH-mutant gliomas versus wild-type samples (n=21 versus 30 patients), and **(D)** lumbar CSF from recurrent IDH-mutant gliomas versus wild-type samples (n=9 versus 18 patients). **(E)** D-2-HG concentrations were compared by Mann Whitney U tests in IDH mutant versus wild-type samples obtained from subarachnoid (n=30 patients per group) or ventricular (n=12 versus 23 patients) CSF, resection cavity (n=10 patients per group), or cyst fluid (n=7 versus 4 patients).

We then evaluated CSF D-2-HG in recurrent IDH-mutant gliomas as a secondary cohort. Intracranially, CSF D-2-HG could distinguish recurrent IDH-mutant gliomas from IDH-wild type samples with an AUC of 0.97 (n=21 versus 30 patients; 95% CI=0.93 to 1.00) (**Figure 2Ci**). Sensitivity and specificity were 96.67% and 90.48% at a cut-off of a D-2-HG concentration of 0.53 μM. Aligned with this, CSF D-2-HG was significantly higher in IDH-mutant gliomas than IDH-wild type samples (median IDH-mutant=3.73 μM, IDH-wild type=0.25 μM; p<0.0001) (**Figure 2Cii**). However, as with primary lesions, lumbar CSF D-2-HG could not distinguish recurrent IDH-mutant gliomas from IDH-wild type samples in ROC analyses (AUC=0.60, 95% confidence interval=0.38-0.83) (**Figure 2Di**) nor in Mann Whitney U-tests (median IDH-mutant=0.20 μM, IDH-wild type=0.19 μM; p=0.3960) (**Figure 2Dii**). Finally, given variations in cranial CSF D-2-HG across compartments, we evaluated the impact of anatomic location on CSF D-2-HG differences between IDH-mutant gliomas and IDH-wild type brain tumors and non-tumor pathologies. Across all cranial compartments, CSF D-2-HG was higher in IDH-mutant gliomas than wild-type samples (**Figure 2E**). The greatest overlap between IDH-mutant and IDH-wild type CSF samples was identified in subarachnoid CSF compared to ventricular CSF and resection cavity or cyst fluid. Indeed, in the two most well-powered compartments, we observed 76.67% sensitivity and 96.67% specificity for IDH-mutant gliomas in subarachnoid CSF at a cut-off of 0.4682 μM (n=30 patients; **Supplementary Figure 5A**) compared to 100% sensitivity/specificity at a cut-off of 0.4760 μM in ventricular CSF (n=12 IDH-mutant vs. n=23 wild-type; **Supplementary Figure 5B**). More direct tumor-CSF contact in the latter three compartments may account for these differences, although these compartments were not as well-powered as subarachnoid CSF. Differences in D-2-HG across compartments were not present in IDH-wild type samples. In conclusion, intracranial, but not lumbar, CSF can be used to distinguish IDH-mutant gliomas from IDH-wild type brain tumors and non-tumor pathologies.

### Longitudinal cranial CSF D-2-HG during cytoreduction

As intracranial CSF D-2-HG is significantly elevated in IDH-mutant gliomas, we hypothesized that this analyte may be responsive to changes in disease burden when sampled serially from CSF access devices like Ommaya reservoirs or ventriculoperitoneal (VP) shunts. In seventeen patients with pre-versus-post-resection/pre-chemoradiation CSF, D-2-HG decreased to a median of 0.54x of its baseline level (range=0.01-1.96x; p=0.0129 by Wilcoxon signed-rank test; median post-operative day=16, range=1-91 days) (**Figure 3Ai**). Extent of resection did not impact the magnitude of decrease in CSF D-2-HG (R^2^=0.0892; **Supplementary Figure 4B**). This may reflect the impact of surgery on local anatomy hampering paired sampling from identical locations before and after surgery. In the three patients for whom CSF D-2-HG increased after resection, an intra-parenchymal catheter in patient 265 (likely sampling interstitial fluid rather than CSF) (**Figure 3Aii**) and greater residual tumor-CSF contact in patients 416 and 426 (**Figure 3Aiii-iv)** may explain this finding.

**Figure 3.**
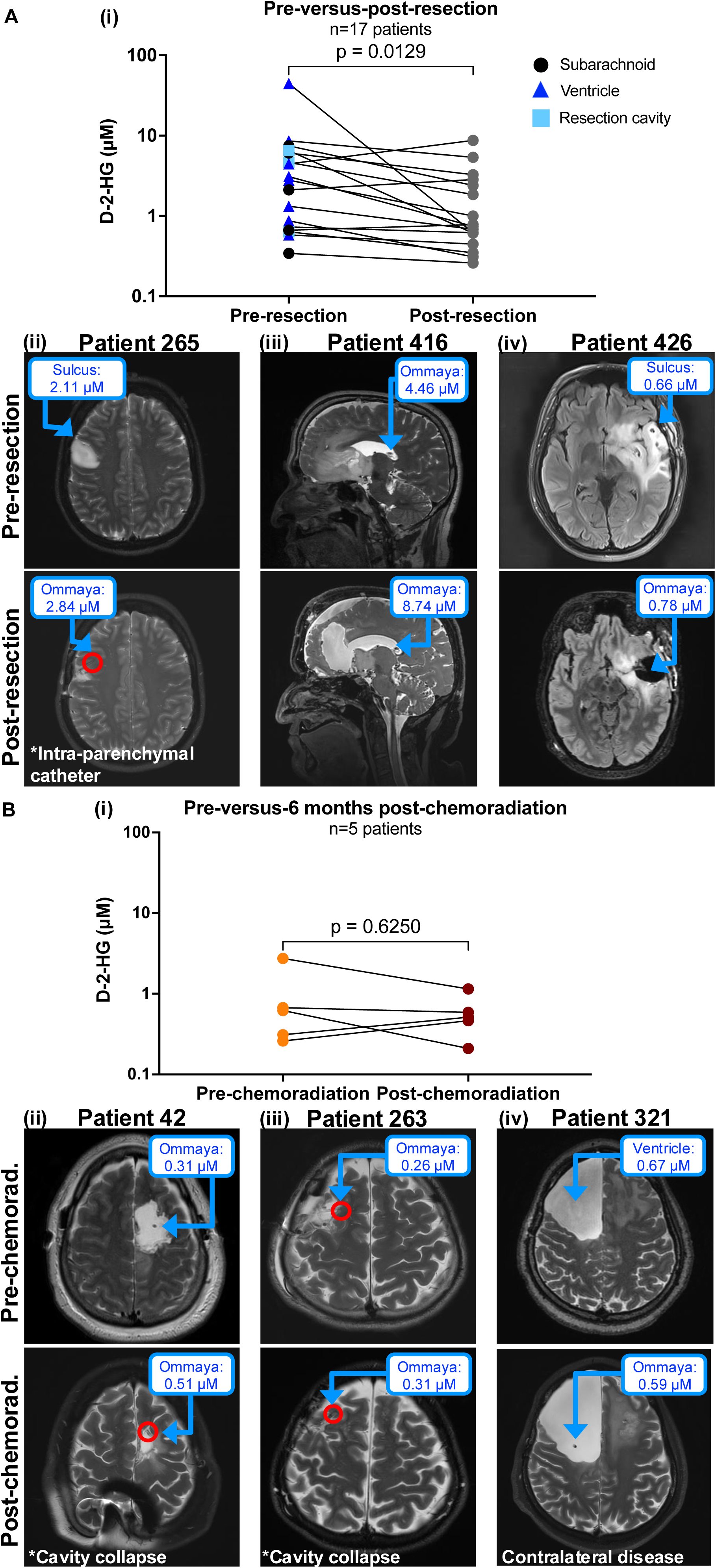
CSF D-2-HG was evaluated prior to versus after cytoreduction. (A) **(i)** A Wilcoxon signed-rank test was performed on CSF samples obtained pre-versus-post-resection, pre-chemoradiation from seventeen patients with IDH-mutant gliomas. Pre-versus-post-resection MRIs and the associated D-2-HG concentrations are shown for the three patients for whom CSF D-2-HG increased with resection **(ii)** patient 265, **(iii)** patient 416, and **(iv)** patient 426. **(B) (i)** A Wilcoxon signed-rank test was performed on CSF samples obtained prior to chemoradiation versus at least six months after the end of chemoradiation in five patients with IDH-mutant gliomas. The pre-versus-post-chemoradiation MRIs and the associated D-2-HG concentrations are shown for the three patients for whom CSF D-2-HG did not decrease substantially with chemoradiation, **(ii)** patient 42, **(iii)** patient 263, and **(iv)** patient 321. The two patients for whom D-2-HG did decrease substantially were patient 46 (Fig. 4A) and patient 245 (Fig. 4D).

As most patients with IDH-mutant high-grade gliomas undergo chemoradiation, we then asked whether CSF D-2-HG decreases with these treatments. D-2-HG was compared between CSF post-resection/pre-chemoradiation and 6 months post-chemoradiation to capture cell death that can occur after radiation ends. CSF D-2-HG decreased substantially in only 2/5 patients (**Figure 3Bi**), patients 46 and 245 – both of whom had open resection cavities with ventricular contiguity. In two of the patients where CSF D-2-HG increased with chemoradiation, the resection cavity had collapsed on the catheter (**Figure 3Bii-iii**), leading to sampling of interstitial parenchymal fluid rather than CSF. The third patient had recurrent disease at time of Ommaya reservoir placement, with Ommaya reservoir placed in the original resection cavity contralateral to the new recurrence (**Figure 3Biv**). Unfortunately, the recurrent disease demonstrated no radiographic response to therapy. As such, documentation of declining D-2-HG with therapy requires a catheter that is not within a collapsed cavity, and a tumor that is responding to therapy administered.

### Disease monitoring in patients with grade 4 IDH-mutant astrocytomas

Ultimately, we aim to assess if CSF D-2-HG can complement disease monitoring, particularly in cases equivocal for recurrence versus treatment-related effects. Thankfully, few patients to date in our series have experienced overt recurrences, although we have recently reported early results of increasing CSF D-2-HG in one patient experiencing radiographic disease recurrence^12^. We also previously presented the initial disease courses of our first three patients with grade 4 IDH-mutant gliomas for whom serial CSF had been acquired, patients 46, 98, and 138^9^. To demonstrate how CSF D-2-HG is being evaluated within patients, we herein present these three patients’ updated CSF D-2-HG data with over two years of follow up, as well as that of the two additional patients with grade 4 IDH-mutant astrocytomas.

Since our original pilot study report^9^, patient 46 has remained stable with no radiographic evidence of disease recurrence nearly 3 years after surgery. Consistent with this, CSF D-2-HG has remained stably low since the end of adjuvant TMZ at a median concentration of 0.19 μM during surveillance (range: 0.13-0.25 μM; CV=23.93%) (**Figure 4A**). In patient 98, further follow-up after two additional years confirmed our initial suspicion of pseudoprogression based on continuing decrease in CSF D-2-HG, as evidenced by decreasing contrast-enhancement since POD305 (**Figure 4B**). Around POD532, a small new contrast-enhancing nodule not in direct contact with the resection cavity prompted initiation of temozolomide with a PARP inhibitor. No increase was seen in D-2-HG at that time, likely due to minimal tumor-CSF contact at the nodule. This nodule has since regressed, and patient has remained stable for another 14 months with a stable median D-2-HG of 0.30 μM (range: 0.26-0.45 μM; CV=22.48%). In patient 138, after the previously reported decrease in D-2-HG with resection, disease progression was suspected soon after completion of adjuvant TMZ based on new periventricular enhancement. CCNU was initiated and all six cycles completed, with disease stability for over four months since then. While CSF D-2-HG increased by 2.49x to 1.00 μM at suspected progression, it stabilized while on CCNU and slightly decreased to a median of 0.60 μM (range: 0.52-0.63 μM; CV=9.74%) during surveillance thereafter (**Figure 4C**).

**Figure 4.**
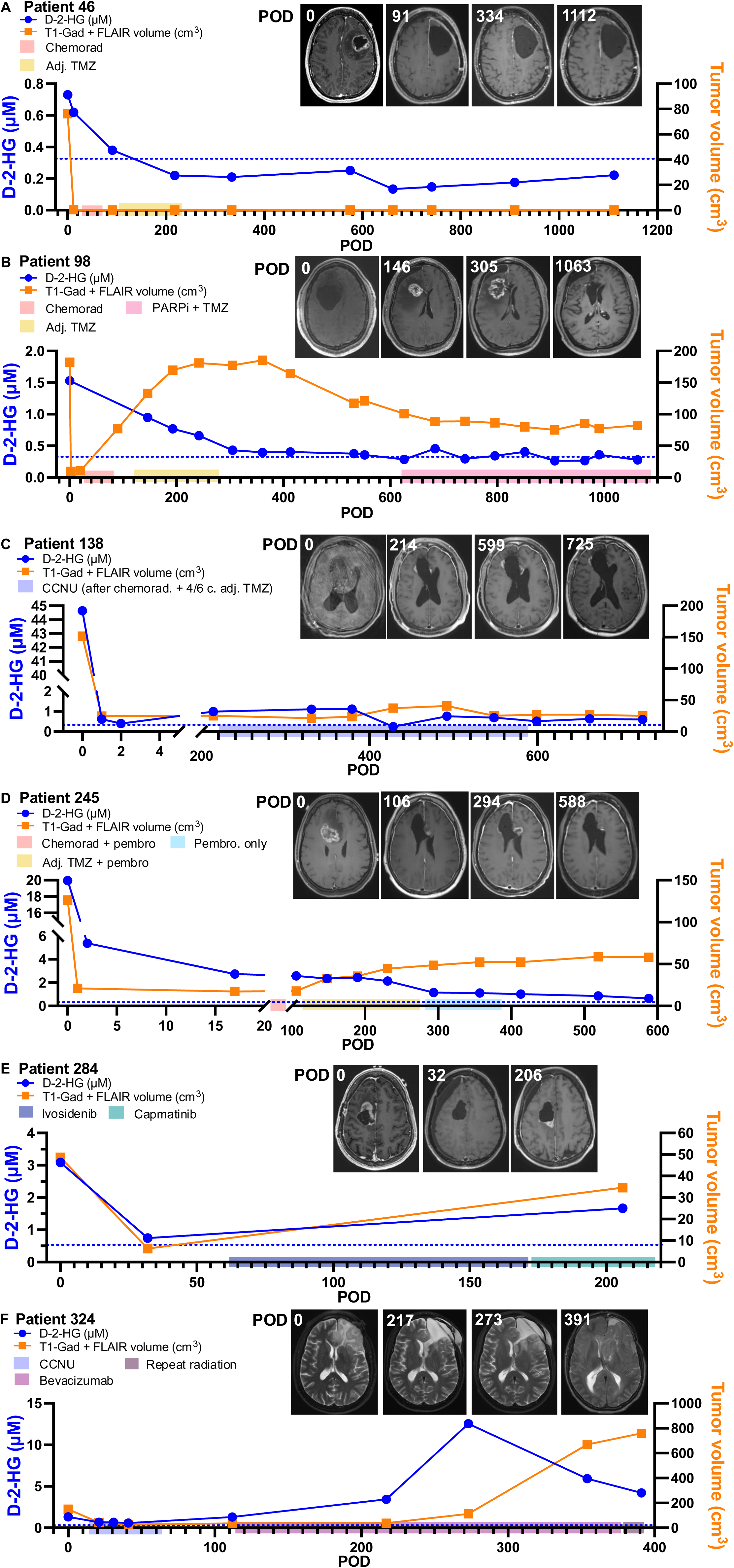
CSF D-2-HG was evaluated in serial samples from patients with grade 4 IDH-mutant astrocytomas. CSF D-2-HG concentrations were evaluated in serial CSF samples obtained from six patients with grade 4 IDH-mutant astrocytomas, as well as total tumor volume from MRIs (in cm^3^, T1-Gadolinium and T2-FLAIR), including **(A)** patient 46, **(B)** patient 98, **(C)** patient 138, **(D)** patient 245, **(E)** patient 284, and **(F)** patient 324. Different treatments ongoing at the time of sample collection are indicated by colored bars on the x-axis. Blue dotted line in all cases = 0.33 μM, indicative of cut-off for primary IDH-mutant gliomas from Figure 2A(i).

In addition to these three patients with grade 4 IDH-mutant astrocytomas, serial CSF samples have also been acquired for over a year from another patient with a grade 4 IDH-mutant astrocytoma, patient 245. By POD17, CSF D-2-HG obtained from an Ommaya reservoir fell to 2.74 μM, or 0.14x of baseline intra-operative levels (**Figure 4D**). D-2-HG continued to decrease after chemoradiation and adjuvant TMZ to a median of 1.11 μM during surveillance (range: 1.02-1.15 μM; CV=5.90%). Similarly to patient 98, increased contrast-enhancement was seen during and after chemoradiation that resolved by POD413, consistent with treatment-related effects and decreasing CSF D-2-HG concentrations.

CSF was also serially sampled for patient 284, who underwent surgery for a recurrent grade 4 IDH-mutant astrocytoma. CSF D-2-HG decreased to 0.24x of baseline levels after resection, and increased thereafter by 2.25x with clear radiographic progression posterior to the resection cavity while on capmatinib (**Figure 4E**). Although no CSF samples were obtained, the patient further progressed through capmatinib and one cycle of lomustine, before transitioning to bevacizumab and passing away on POD497. Finally, in patient 324 who also had a grade 4 IDH-mutant astrocytoma, CSF D-2-HG increased by prior to clear disease progression. However, after POD354, CSF D-2-HG decreased substantially, which was incongruent with radiographic progression (**Figure 4F**). Review of imaging revealed that anatomic changes had led to obstruction of CSF flow and sampling, resulting in inadequate sampling of CSF. In summary, when there is adequate tumor-to-CSF contact, CSF D-2-HG may be useful for delineating true disease progression from treatment-related effects in grade 4 IDH-mutant astrocytomas.

## DISCUSSION

Improved methods for longitudinal disease monitoring are needed for patients with gliomas^14^. In evaluating CSF D-2-HG as a biomarker for IDH-mutant gliomas, our study revealed significant differences between lumbar and cranial CSF D-2-HG concentrations, with low lumbar levels precluding the utility of CSF D-2-HG as a diagnostic or monitoring biomarker. Notably, cranial CSF D-2-HG concentrations reflected changes in disease burden, though access to an anatomically stable non-collapsed CSF compartment is required for reliable monitoring. Therefore, with optimal catheter placement and minimal resection cavity changes, longitudinal CSF D-2-HG may reflect therapeutic response and aid in distinguishing recurrence from treatment-related imaging effects.

D-2-HG concentration gradients between intracranial and lumbar CSF in patients with IDH-mutant gliomas have previously been reported by our group using 14 paired samples^12^ and by Kalinina et al. with unpaired samples from 3 lumbar and 41 intracranial CSFs^10^. Our updated analysis, including 15 and 42 patients with lumbar and intracranial CSF, respectively, again demonstrate a neuroaxis gradient for D-2-HG. However, unlike our study, Kalinina et al. reported that lumbar CSF D-2-HG could distinguish IDH-mutant gliomas from IDH-wild type controls with 100% sensitivity and specificity^10^, albeit with sample sizes of n=3 versus 12 patients, respectively. Notably, the lumbar D-2-HG concentrations in their 3 IDH-mutant gliomas were 0.446, 1, and 20.3 μM, (mean=7.25 μM, SD: 11.31 μM) compared to a mean of 0.19 μM (SD: 0.07 μM) in 12 IDH-wild type controls. In contrast, our data show that lumbar CSF poorly distinguishes IDH-mutant gliomas (n=12; mean=0.18 μM, SD=0.09 μM) from IDH-wild type controls (n=13; 0.1601 μM, SD: 0.05 μM) (**Figure 2Bi**). Such exemplar cases of elevated lumbar D-2-HG have yet to be observed in our practice (0/22 cases to date). Considering the absence of clinical context available for the de-identified samples provided to Kalinina et al., one must wonder if the lumbar CSF samples were obtained in those patients due to clinical concern for leptomeningeal gliomatosis, although the clinical context for these LPs was not provided in their annotations. To mitigate the challenge of comparing datasets across institutions, we and others have previously encouraged publication of patient-level clinical and technical meta-data^12^, which are provided in Supplemental Data for this work.

While lumbar CSF D-2-HG does not currently appear feasible for diagnostic or monitoring purposes, D-2-HG was significantly higher in cranial IDH-mutant gliomas than IDH-wild type controls. Similarly, Kalinina et al. reported sensitivity and specificity of 63% and 94% in subarachnoid (cisternal) CSF and 82% and 96%, respectively, in ventricular CSF for IDH-mutant gliomas versus IDH-wild type controls^10^. Our findings similar suggest improved sensitivity/specificity for discriminating IDH-mutant gliomas from IDH-wild type controls in ventricular (100% sensitivity/specificity) than subarachnoid CSF (76.67% sensitivity/96.67% specificity) (**Supplementary Figure 5**). However, this finding is biased by greater tumor-to-CSF contact in the ventricular group, as a resection that reached the ventricle would have involved tumor at the ventricular surface. Both their study and ours demonstrate marked variation in CSF D-2-HG between patients with IDH-mutant gliomas, consistent with previous work in IDH-mutant cell lines^13^. While the diagnostic relevance of cranial CSF is limited, considering it is typically initially accessed only at the time of tissue sampling, intra-operatively obtained CSF results have provided early evidence of an IDH mutation in cases that were negative for IDH1-R32H immunohistochemistry, and for which sequencing results were not yet available. Matrix-assisted laser desorption ionization (MALDI) and desorption electrospray ionization (DSI) mass spectrometry imaging has also been used for rapid intra-operative determination of D-2-HG that could help surgeons tailor extent of resection^15–18^. While it cannot provide spatial information, GCMS is more widely available than MALDI. Our CLIA test could enable intra-operative diagnosis to guide intra-operative decision-making. Indeed, resection of non-enhancing disease has been reported to have greater prognostic impact in IDH-mutant than wild-type astrocytoma^19,20^.

Monitoring data demonstrate the ability to utilize serial CSF D-2-HG to confirm disease response despite treatment-related changes, as demonstrated in patients 98 and 245 (**Figure 4B,D**), and to confirm disease progression in patients 284 and 324 (**Figure 4E,F)**. These patients, along with two prior patients in Fujita et al.^11^, underscore the potential of CSF D-2-HG for disease monitoring. However, the lack of substantial decrease in CSF D-2-HG in 2/5 patients six months after chemoradiation underscores the need for attention to detail, including clinical trajectory (for example, radiation-resistant recurrence in patient 321) and anatomy. If the anatomy of the regions being compared is no longer comparable, as in patients 42 and 263, results may also be incomparable. Collapse of the resection cavity onto the catheter can lead to sampling of low volumes of interstitial fluid rather than higher volumes of CSF^12^, artificially increasing serial CSF D-2-HG concentrations. Indeed, the patients for whom CSF D-2-HG correlated best with disease trajectory were those with open resection cavities that remained in continuity with the ventricular system (**Figure 4**). Even in these patients, tumor-to-CSF contact needs to be evaluated if there is concern for disease progression in an area with no CSF contact, as demonstrated by patients 98 and 324. In some patients, the relative burden of recurrence versus latent disease may also prevent early detection of recurrence. Indeed, in patient 98, CSF cell-free DNA also did not increase with suspected progression^21^; however, no tissue was collected to confirm or refute recurrence. Finally, for patients with no radiographically detectable tumor post-resection, we anticipated intracranial CSF D-2-HG levels similar to those of IDH-wild type patients; however, levels sometimes remained above the cut-off of 0.33 μM identified for primary IDH-mutant gliomas, which could open the door to monitoring impact of therapies aiming to further ablate residual disease^22^. Further work will be needed to determine optimal thresholds for identifying therapeutic response or disease progression via CSF D-2-HG. Deviation of CSF D-2-HG outside of the coefficient of variance during stable disease may be an option, as previously described for tumor growth rates^23–25^.

Our study has several limitations including its single-center design and the absence of an external validation cohort. However, key findings of elevated cranial CSF D-2-HG in IDH-mutant gliomas was separately demonstrated in two separate cohorts: primary and recurrent gliomas, with elevations observed in CSF from multiple intracranial compartments. Tissue D-2-HG levels were not measured to assess for correlation to CSF. Although Kalinina et al.^10^ previously observed correlation between tissue and cranial CSF levels, impacts of proximity to the sampled CSF compartment would still be expected. Although tumor-to-CSF contact is hypothesized to influence CSF D-2-HG levels, standardized methods for quantifying tumor-to-CSF surface area and D-2-HG diffusion to CSF would need to be developed. Finally, in patients with question of disease progression, tissue was not obtained to confirm or refute disease progression. This lack of tissue sampling means that no gold-standard is available against which to benchmarking of CSF D-2-HG findings^26^, similar to challenges encountered in our prior cell-free DNA studies. Although survival can be used as a proxy, some patients with recurrent IDH-mutant may respond to therapeutic re-challenge, limiting the utility of survival for assessing progression versus pseudoprogression. In some cases, comparison to other tumor-associated candidate CSF biomarkers may help with cross-validating CSF 2-HG results during disease monitoring^9,12^. Indeed, as IDH inhibitors become routine for patients with low-grade IDH-mutant gliomas, candidate CSF biomarkers other than D-2-HG, such cell-free DNA^21,27,28^ or proteomics^29–31^, will likely need to be evaluated as IDH inhibition decreases D-2-HG concentrations. In conclusion, with future validation in larger cohorts, we hope that CSF D-2-HG may be leverageable alongside MRIs and other CSF biomarkers for monitoring of disease progression versus pseudoprogression in patients with IDH-mutant gliomas.

## Authorship

Conceptualization: CRC, PAD, JEP, SHK, TJK, TCB

Sample acquisition and processing: CRC, AEW, TCB

Performing experiments: CRC, YS, ZK, JML, WJL

Data analysis: CRC, YS, ZK, JML, WJL, TCB

Protocol and project administration: CRC, TCB

Writing – original draft: CRC, TCB

Writing – review & editing: all authors

## Data availability

All data are available as supplementary files. Any other data can be requested from the corresponding author.

## Acknowledgments

We thank our patients and their families for their participation in this study. We thank the Mayo Clinic Neurosurgery Clinical Research, neurosurgical residents, fellows, and midlevels, and surgical support and clinical research staff for their invaluable contributions to this work.

## Ethics

This study was approved by the Mayo Clinic Institutional Review Board and all participants provided their consent to participate in this study. This study was performed in accordance with the Declaration of Helsinki.

## Consent

All participants have provided consent for publication.

