## Supplemental Methods, Note, Figures, and table for "Cerebrospinal fluid D-2-hydroxyglutarate for IDH-mutant glioma: utility for detection versus monitoring"

**Supplementary Materials**

**Supplementary Methods**

*CLIA validation of CSF D/L-2-HG assay (Test ID: 2HGCF)*

To meet CLIA guidelines for clinical deployment of the assay, intra-and-inter-assay precision were evaluated, with pre-determined acceptance criteria of a technical coefficient of variance (CV) of ≤ 20%. Intra-assay precision was measured by evaluating three different CSF levels twenty times each in a single batch, using CSF spiked with 1, 10, or 20 μM of D or L-2-HG. Inter-assay precision was measured by evaluating these three CSF levels across twenty batches. Accuracy was evaluated by spiking three waste specimens at 2.5 or 5 μM of D-or-L-2-HG and calculating recovery as (final concentration-baseline concentration)/added concentration, x 100%. Percent recovery was averaged across three specimens, with acceptance criterion set at an average recovery of 100% ± 20%. Reportable range was evaluated by measuring the lower limit of quantification (LLOQ) and upper limit of quantification (ULOQ) in artificial CSF. The LLOQ was identified as the concentration at which a signal to noise (S/N) of 10.1 was reached, using the built-in S/N macro on the Agilent ® ChemStation. This functions by measuring the signal intensity in a blank sample (known as the zero calibrator) by analyzing the extracted ion chromatogram at the specific mass-to-charge ratio that corresponds to the compound being quantified. The measurement is performed at the expected retention time of the compound in the chromatogram and includes assessing the background noise near that retention time. The ULOQ was identified by identifying the maximum concentration of D-or-L-2-HG at which linearity was maintained according to a coefficient of determination (R^2^) ≥ 0.98, evaluated in three separate batches. Linearity within the analytical measurement range (AMR) was determined by diluting the high control with CLR water at x2, x4, and x8. The acceptance criterion was that the percent difference between measured and expected values were 0 ± 25%, with a slope between the measured and expected of 0.9-1-1 and a coefficient of determination (R^2^) ≥ 98%. To evaluate linearity above the analytical measurement range, an artificial CSF specimen spiked at 200 μM was serially diluted with water to the LLOQ. The acceptance criterion was identical to that of linearity within the AMR. Analytical specificity was evaluated by spiking an aliquot of CSF with 15 or 30 μM of 3-OH glutaric acid as the metabolite with the same molecular weight as D-and-L-2-HG. The percent change between the baseline and spiked specimen was calculated as (spiked-baseline sample)/baseline sample x 100%, with an acceptance criterion of 0% ± 20%.

*D-and-L-2-HG stability experiments*

To determine the stability of D-or-L-2-HG in CSF at different temperatures, a CSF specimen from an external ventricular drain (EVD, IDH-WT) was thawed and spiked with 5 μM of D-or L-2-HG. Specimens were then stored at -80°C, 20°C, or 37°C for 24 hours before analysis. To evaluate the impact of blood or plasma contamination, whole blood was obtained from a donor. Plasma was generated by double-spinning this whole blood at 2,000xG for 10 minutes at 4°C. The CSF was then spiked to 5% whole blood or plasma then immediately spun down at 400xG for 10 minutes at 4°C, and supernatant collected. Then, the supernatant was spiked with 5 μM of D-or-L-2-HG at room temperature and then stored at -80°C before analysis. Finally, to evaluate whether antibiotics present within the Bactiseal catheter, or utilized locally during Ommaya placement could interfere with the assay, CSF spiked with 5 μM D- or L-2-HG was also spiked with supratherapeutic levels (250 μg/m) each of vancomycin, gentamicin, rifampin, and clindamycin at room temperature and then stored at -80°C before analysis.

**Supplementary Note**

*Developing the CLIA-level assay for CSF D-and-L-2-HG*

To be used for clinical decision-making, assays should meet Clinical Laboratory Improvement Amendments (CLIA) criteria . As such, in collaboration with the CLIA-certified Mayo Clinic Biochemical Genetics Laboratory, we repurposed an existing gas chromatography-mass spectrometry (GCMS) D-and-L-2-HG assay originally developed for urine. Analyses were performed using the CSF matrix to assess performance based on Clinical and Laboratory Standards Institute (CLSI) guidelines for inter-and-intra-assay precision (technical coefficient of variance), accuracy, reportable range and linearity, and analytical specificity. CSF samples were spiked with 1, 10, or 20 μM each of D-and-L-2-HG and analyzed twenty times in the same run. The technical coefficient of variance (CV) ranged from 2-5% for D-or-L-2-HG across the three different D-and-L-2-HG concentrations (**Supplementary Figure 1A**). These same CSF samples were then analyzed in twenty different runs and had a CV ranging from 2-10% (**Supplementary Figure 1B**). In sum, intra-and-inter-assay precision were both acceptable per the acceptance criteria (≤ 20% CV).

Accuracy was then verified from recovery of CSF specimens spiked with 2.5 or 5 μM each of D-and-L-2-HG in triplicate (**Supplementary Figure 1C**). At 2.5 μM, the average D-and-L-2-HG recoveries were 113% and 105%, respectively. At 5 μM, the accuracy was 108% for both D-and-L-2-HG. The acceptance criterion of 100% ± 20% was met at both concentration levels. The reportable range of D-and-L-2-HG was then assessed by identifying the lower limit of quantification (signal to noise = 10.1) and upper limit of quantification (maximum concentration with coefficient of determination, R^2^≥ 0.98). Concentrations of 0.0010 μM for D-2-HG and 0.0005 μM for L-2-HG resulted in a signal to noise ratio of 10.1. For D-2-HG, a maximum concentration of 150 μM evaluated three separate times resulted in R^2^ values of 0.9990, 0.9994, and 0.9984; a maximum concentration of 150 μM for L-2-HG resulted in R^2^ values of 0.9991, 0.9995, and 0.9983. The analytical measurement range for D-2-HG was 0.0010 to 150.00 μM and 0.0005 to 150.00 μM for L-2-HG (**Supplementary Figure 1D; Supplementary Data**). Finally, analytical specificity, or assay interference, was evaluated by spiking in 15 or 30 μM of 3-OH glutaric acid, a metabolite with the same molecular weight as D-and-L-2-HG. The percent change in D-2-HG between baseline and spiked samples ranged from -15% to 8% (**Supplementary Figure 1E**), meeting the acceptance criterion of an absolute percent change in quantification less than 20% relative to baseline. The linearities of the CSF calibrations are found in **Supplemental Table 1**. In conclusion, all CLIA-level, pre-established acceptance criteria were met for the GCMS CSF D-and-L-2-HG assay, enabling its availability as a clinical test at our institution.

*Impact of temperature, blood/plasma contamination, and antibiotics on D-and-L-2-HG quantification in CSF*

The stability of D-2-HG in CSF as a biologically relevant matrix has only been assessed with freeze-thawing and exposure to room temperature. CSF can be exposed to varying temperatures, blood or plasma contamination, and antibiotics that could impact analyte stability or assay interference. As such, independent of the CLIA criteria above, we evaluated the impact of temperature on spiked D-and-L-2HG in CSF at three different temperatures based on relevant real-world scenarios: (1) -80°C, the temperature at which samples are biobanked, (2) 20°C, replicating prolonged CSF exposure to room temperature if there samples are not kept on wet ice during delays between collection and processing, and (3) 37°C, the physiologic body temperature to which CSF is exposed before collection at surgery or from CSF access devices. At all temperatures over 24 hours, there were no statistically significant differences in measured D-or-L-2-HG concentrations (**Supplementary Figure 1Fi-ii**). Then, CSF samples were spiked with D-and-L-2-HG and either 5% blood or plasma to mimic blood or plasma contamination that can occur during sample collection or from a disrupted blood-brain barrier, respectively. D-and-L-2-HG concentrations were not significantly different than that of the control CSF sample not spiked with plasma or blood (**Supplementary Figure 1Gi-ii**). Finally, in our clinical protocols, intracranial CSF may be exposed to antibiotic-eluting ventricular catheters as well as antibiotic solution injected into the reservoir. To assess potential assay interference, we analyzed CSF spiked with D- and L-2-HG, with and without the addition of 250 μg/mL of each antibiotic, and observed no significant differences in measured concentrations (**Supplementary Figure 1Hi-ii**). In summary, D- and L-2-HG were minimally impacted by exposure to varying temperatures, blood or plasma contamination, and antibiotics.

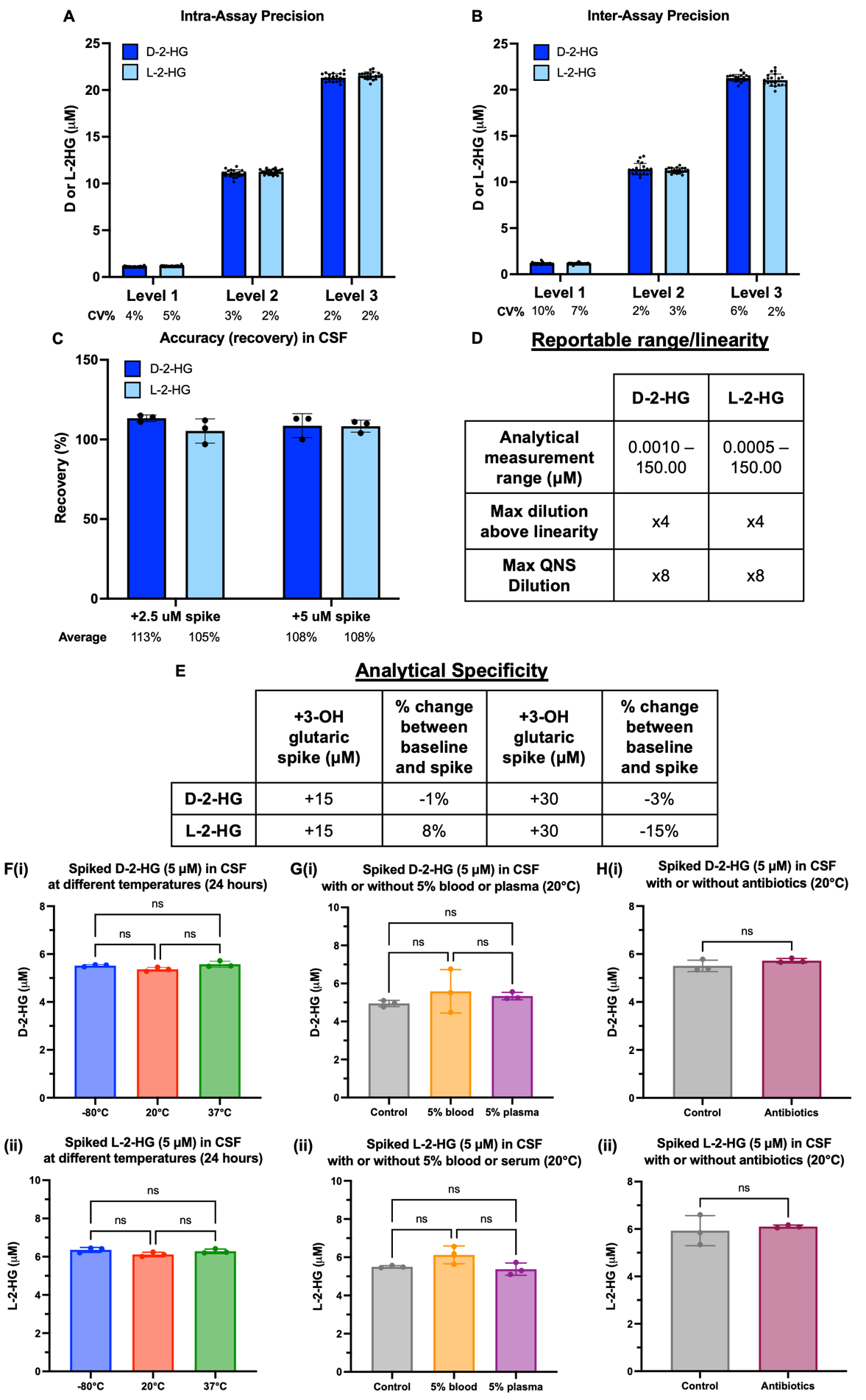
**Supplementary Figures and Figure Legends**

**Supplementary Figure 1. CLIA validation of CSF D/L-2-HG assay and stability of CSF D or L-2-HG. (A)** Intra-assay precision was assessed based on measurement of D or L-2-HG concentrations after CSF was spiked at 1, 10, or 20 μM. Twenty measurements were taken in each batch. The coefficient of variance for D or L-2-HG was calculated at each spike level. **(B)** Inter-assay precision was assessed by measuring D-or-L-2-HG concentrations across twenty run after spiking CSF at 1, 10, or 20 μM. The coefficient of variance for D or L-2-HG was calculated at each spike level. **(C)** Accuracy, or recovery, of CSF D-or-L-2-HG was measured at two different spiked concentrations (2.5 or 5 μM), with 3 replicates at each spike level. **(D)** Reportable range and linearity are summarized for both D and L-2-HG based on the analytical measurement range (AMR), maximum dilution above linearity, and maximum dilution within the AMR (QNS = quantity not sufficient). **(E)** The analytical specificity of the assay is reported based on spiking 15 or 30 μM of 3-OH glutaric acid in CSF and measuring D or L-2-HG. **(F)(i)** D-2-HG or **(ii)** L-2-HG concentrations were measured after D or L-2-HG was spiked into CSF at 5 μM and exposed to three different temperatures (-80°C, 20°C, or 37°C) for 24 hours. **(G)** **(i)** D-2-HG or **(ii)** L-2-HG concentrations were measured after whole blood or plasma were spiked to 5% each and then spun down. The supernatant was then spiked with D or L-2-HG at 5 μM before measurement. **(H)(i)** D-2-HG or **(ii)** L-2-HG concentrations were measured after D-or-L-2HG at 5 μM and supratherapeutic concentrations of antibiotics were spiked into CSF.

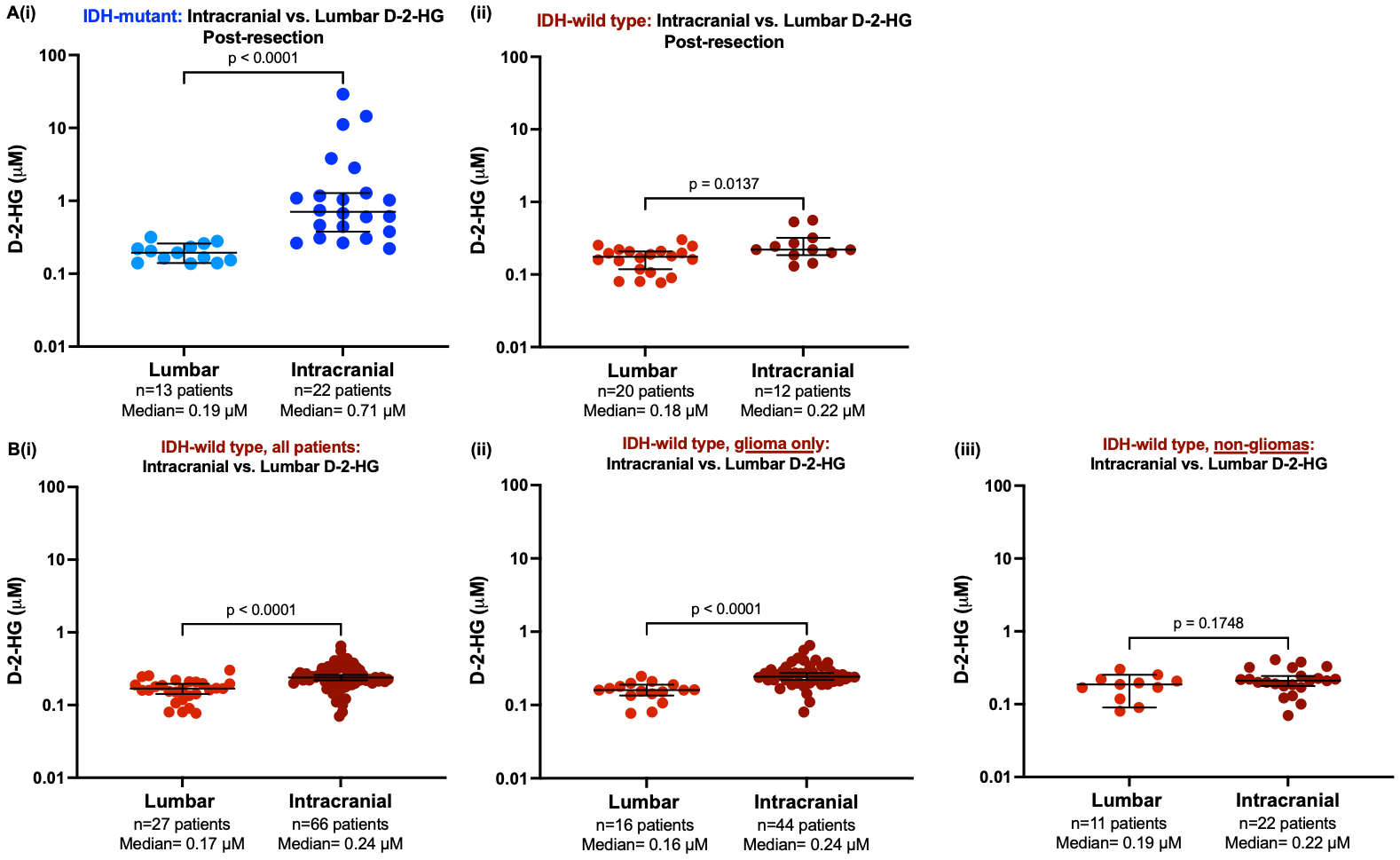

**Supplementary Figure 2**. **Cranial versus lumbar CSF D-2-HG. (A)** CSF D-2-HG concentrations were compared by Mann Whitney U tests in cranial versus lumbar CSF obtained after resection in **(i)** patients with IDH-mutant gliomas (n=22 cranial, 13 lumbar) or **(ii)** IDH-wild type controls (n=12 cranial, 20 lumbar). (**B)** Mann-Whitney U-tests were used to compare CSF D-2-HG concentrations in cranial versus lumbar CSF in **(i)** all IDH-wild type controls (n=66 cranial, 27 lumbar), **(ii)** only IDH-wild type glioma (n=44 cranial, 16 lumbar), and **(iii)** non-glioma IDH-wild type controls (n=22 cranial, 11 lumbar). Lines are medians and bars are 95% confidence intervals.

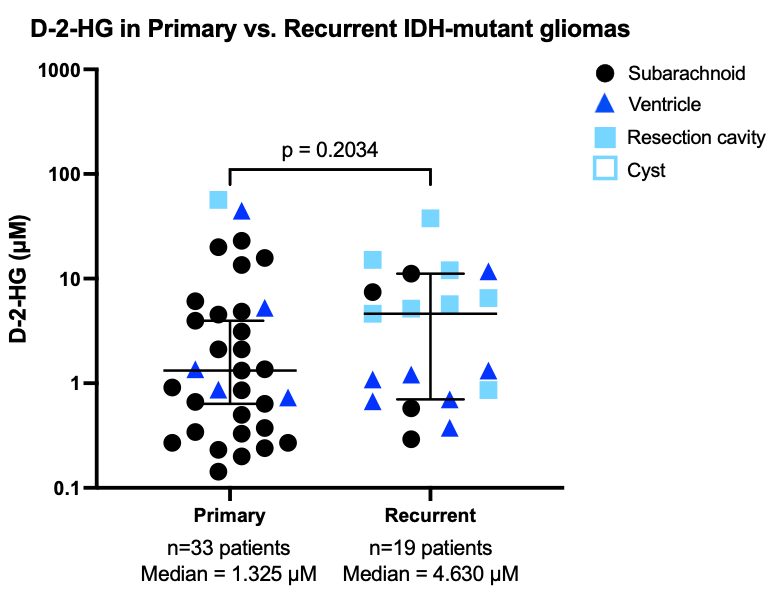

**Supplementary Figure 3**. **Cranial CSF D-2-HG in primary versus recurrent IDH-mutant gliomas.** CSF D-2-HG concentrations were compared by Mann Whitney U-tests in patients with primary (n=33) or recurrent (n=19) IDH-mutant gliomas. Of note, a resection cavity sample was obtained in one primary IDH-mutant glioma patient due to a prior subtotal resection at an outside institution. Location of CSF sampling are indicated by different symbols. Lines are medians and bars are 95% confidence intervals.

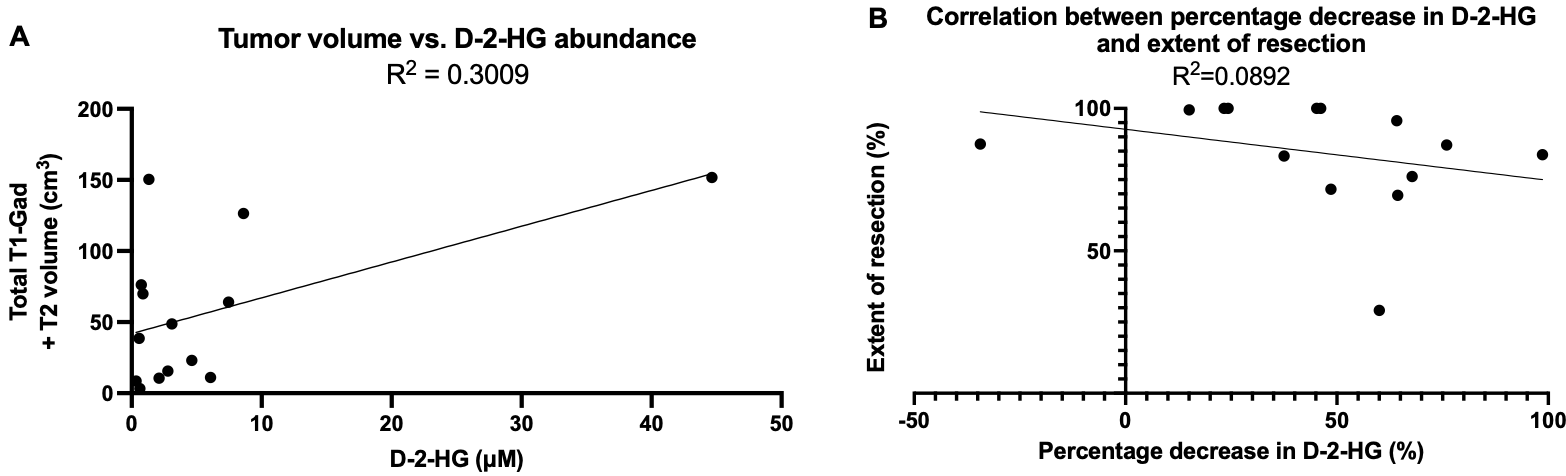

**Supplementary Figure 4. Volumetrics and CSF-D-2-HG. (A)** In patients where volumetrics was obtained prior to tumor resection, CSF D-2-HG concentrations were correlated to tumor volume (in cm^3^, total T1-gadolinium and T2 volume). **(B)** Extent of resection was correlated to the
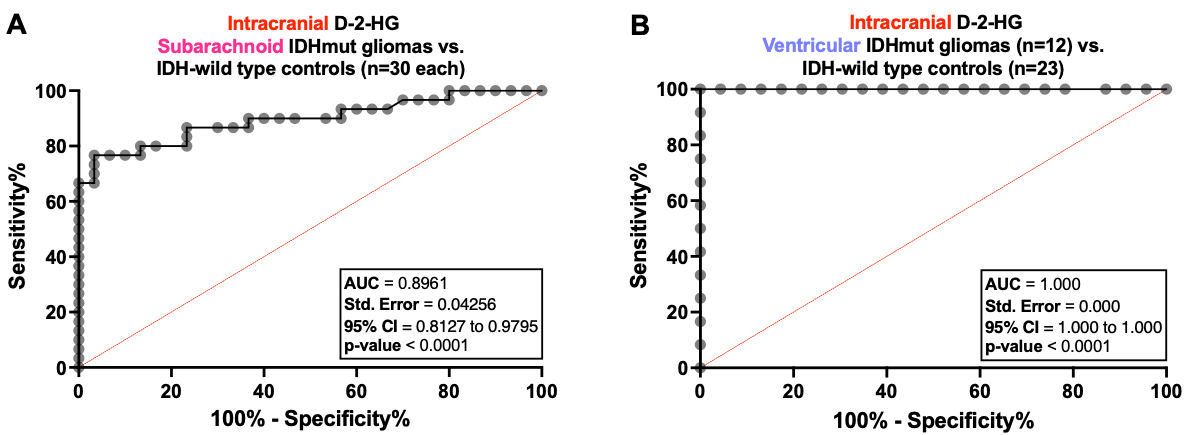
percentage decrease in D-2-HG prior to versus after resection.

**Supplementary Figure 5. IDH-mutant glioma detection in subarachnoid versus ventricular CSF.** Receiver operating characteristic (ROC) analyses were performed to evaluate the sensitivity versus specificity of **(A)** subarachnoid CSF D-2-HG or **(B)** ventricular CSF for distinguishing IDH-mutant gliomas from IDH-wild type controls (subarachnoid CSF: 30 patients each for IDH-mutant gliomas versus wild-type controls; ventricular CSF: 12 IDH-mutant gliomas versus 23 IDH-wild type controls).

**Supplemental Table 1. CSF Calibration Curves run across three different days.**

| A. **11/9/2021** |  |  |
| --- | --- | --- |
| **theoretical ratio** | **L-2HGA** | **D-2HGA** |
| 0.000 | 0.0009 | 0.0011 |
| 0.125 | 0.1485 | 0.1442 |
| 0.250 | 0.2688 | 0.2683 |
| 0.500 | 0.5404 | 0.5466 |
| 1.000 | 1.0929 | 1.1380 |
| 1.500 | 1.5909 | 1.6088 |
| 10.000 | 10.5392 | 10.5446 |
| 15.000 | 14.6234 | 14.6149 |
| **corr (R2)** | **0.9991** | **0.9990** |
| **slope** | **0.9933** | **0.9924** |
| B. **12/1/2021** |  |  |
| **theoretical ratio** | **L-2HGA** | **D-2HGA** |
| 0.000 | 0.0027 | 0.0023 |
| 0.125 | 0.1365 | 0.1412 |
| 0.250 | 0.2804 | 0.2906 |
| 0.500 | 0.5272 | 0.5408 |
| 1.000 | 1.0765 | 1.1351 |
| 1.500 | 1.5701 | 1.6508 |
| 10.000 | 10.4023 | 10.4282 |
| 15.000 | 14.7182 | 14.6883 |
| **corr (R2)** | **0.9995** | **0.9994** |
| **slope** | **0.9947** | **0.9922** |
| C. **12/30/2021** |  |  |
| **theoretical ratio** | **L-2HGA** | **D-2HGA** |
| 0.000 | 0.0007 | 0.0008 |
| 0.125 | 0.1485 | 0.1468 |
| 0.250 | 0.2713 | 0.2815 |
| 0.500 | 0.5472 | 0.5458 |
| 1.000 | 1.1098 | 1.1528 |
| 1.500 | 1.7112 | 1.7039 |
| 10.000 | 10.7084 | 10.6838 |
| 15.000 | 14.4972 | 14.5113 |
| **corr (R2)** | **0.9983** | **0.9984** |
| **slope** | **0.9904** | **0.9899** |
